# The CircaHealth CircaPain study protocol: A longitudinal multi-site study of the chronobiological control of chronic pain

**DOI:** 10.1101/2024.03.22.24304751

**Authors:** Doriana Taccardi, Hailey GM Gowdy, Lesley Singer, Jennifer Daly-Cyr, Amanda M Zacharias, Zihang Lu, Manon Choinière, M Gabrielle Pagé, Nader Ghasemlou

**Affiliations:** Department of Biomedical & Molecular Sciences, Queen’s University; Chronic Pain Network, McMaster University; Department of Public Health Sciences, Queen’s University; Dept of Anesthesioloy and Pain Medecine de l’Université de Montréal; Research Center of the Centre hospitalier de l’Université de Montréal; Dept. of Psychology Research Center of the Centre hospitalier de l’Université de Montréal; Department of Anesthesiology & Perioperative Medicine, Queen’s University; Centre for Neuroscience Studies, Queen’s University

**Keywords:** chronic pain, epidemiology, circadian rhythm, biomarkers

## Abstract

**Introduction:** One in five Canadians lives with chronic pain. Evidence shows that some individuals experience pain that fluctuates in intensity following a circadian (24-hour) rhythm. Endogenous molecular rhythms regulate the function of most physiological processes, neuroimmunology functions that govern pain mechanisms. Addressing chronic pain rhythmicity on a molecular and biopsychosocial level can advance understanding of the disease and identify new treatment/management strategies. Our CircaHealth CircaPain study uses an online survey combined with ecological momentary assessments and bio-sample collection to investigate the circadian control of chronic pain and identify potential biomarkers. Our primary objective is to understand inter-individual variability in pain rhythmicity, by collecting biopsychosocial measures. The secondary objective accounts for seasonal variability and the effect of latitude on rhythmicity.

**Methods and analysis:** Following completion of a baseline questionnaire, participants complete a series of electronic symptom-tracking diaries to rate their pain intensity, negative affect, and fatigue on a 0-10 scale at 8:00, 14:00, and 20:00 daily over 10 days. These measures are repeated at 6- and 12-months post-enrolment to account for potential seasonal changes. Infrastructure is being developed to facilitate the collection of blood samples from subgroups of participants 2 times per day over 24-48 hours to identify rhythmic expression of circulating genes and/or proteins.

**Ethics and dissemination:** Ethical approval for this study was obtained by the Queen’s University Health Sciences and Affiliated Teaching Hospitals Research Ethics Board. Findings will be published in a relevant scientific journal and disseminated at national and international scientific meetings and online webinars. We maintain a website to post updated resources and engage with the community. We employ knowledge mobilization in the form of direct data sharing with participants. This study is funded by the Canadian Institutes of Health Research (CIHR) (grant PJT-497592) and the CIHR Strategy for Patient-Oriented Research (SPOR) Chronic Pain Network (CPN) (grant SCA-145102).

Ethical approval date: 08 March 2024

Estimated start of the study: April 2024

**Strengths and limitations of this study:**

- Data will be collected using self-report questionnaires only, which may lead to random or systematic misreporting.
- The online nature of the study might affect the diversity in our sample (e.g., the representation of rural and/or underprivileged communities).
- Physical distance from research laboratories with specialized equipment for analyses and biobanking storage might affect accessibility, however, this can be overcome by using mailable dried blood spot collection kits as described.
- Questionnaires used in our study have previously been validated in the chronic pain population and used in several languages.
- Uncovering distinct pain rhythmicity patterns and health outcomes associated with rhythmicity may help develop new treatments for different chronic pain conditions tailored to individual circadian rhythms.

## 1. Introduction

Chronic pain (lasting >3 months)(1) affects more than 1 in 5 individuals(2-4) and accounts for 5.9% of total disability-adjusted life years(5). While multidisciplinary treatments are recognized as the gold standard for pain management(6-9), the underlying mechanisms contributing to its chronicity are largely unknown and the few therapeutic options available offer only mild to moderate benefits(10, 11).

Biopsychosocial factors modulate individuals’ pain experience and cause variability in the presentation and evolution of disease, leading to heterogeneity in treatment responses(12-15). A better understanding of how each person experiences pain may aid the development of new individualized strategies for pain management(14, 16-18). A commonly used method to identify temporal variations in self-reported measures is the ecological momentary assessment (EMA)(19, 20). In recent years, intra-individual variability has been identified as an important aspect to consider, rather than conceptualized as noise(21). For example, chronic neuropathic pain (e.g., diabetic neuropathy) is observed to increase over the course of the day(22) while inflammatory pain syndromes (e.g., osteoarthritis) tend to follow an inverse pattern(23). However, these studies have not investigated specifically inter-individual differences within the same condition or examined biopsychosocial components associated with pain rhythmicity.

Chronobiology examines the contribution of time to biological outcomes including daily, lunar, and seasonal changes. Circadian rhythms help to align physiological functions with the external environment via external cues including light and feeding schedules (24). These rhythms are controlled by a set of core clock genes that orchestrate a tightly regulated transcription-translation feedback loop lasting approximately 24 hours(25). The timing of secreted factors (e.g., melatonin, cortisol, and growth hormone) is regulated by the suprachiasmatic nucleus, which communicates with peripheral clocks located in almost all organs and cells in the body. These circulating factors interact with immune cells, resulting in a bidirectional circadian-immune relationship(26). Evidence across species supports the influence of circadian rhythms on biopsychosocial processes and behaviours(27).

Capturing fluctuations in pain intensity and their associated factors (both psychosocial and molecular), as reported by people with chronic pain, is central to a thoughtful clinical assessment of disease. These fluctuations may themselves provide insight into underlying causes and result in a better understanding of how persistent pain is modulated. We use a methodology that includes psychosocial measures and provides a foundation for biological sample collection to identify pain rhythmicity phenotypes and their associated molecular signatures. Our core multidisciplinary team includes clinical researchers, pain clinicians, biostatisticians, basic scientists, and individuals with lived experience. This study will characterize inter-individual differences in pain experience and identify new biomarkers of pain rhythmicity.

## 2. Methods and analysis

### 2.1 Primary outcome

The primary outcome of this study is to identify phenotypes of pain rhythmicity using ecological momentary assessment and determine their association with biopsychosocial profiles in individuals living with chronic pain across Canada.

### 2.2 Secondary outcome

The secondary outcome of this study is to identify characteristics of pain rhythmicity and determine their associations with the geophysical and seasonal environments of participants.

### 2.3 Study Design

A biopsychosocial approach underpins this longitudinal cohort study (figure 1). This study complies with STROBE guidelines(28) and with the principles of the Declaration of Helsinki(29).

**Figure 1.**
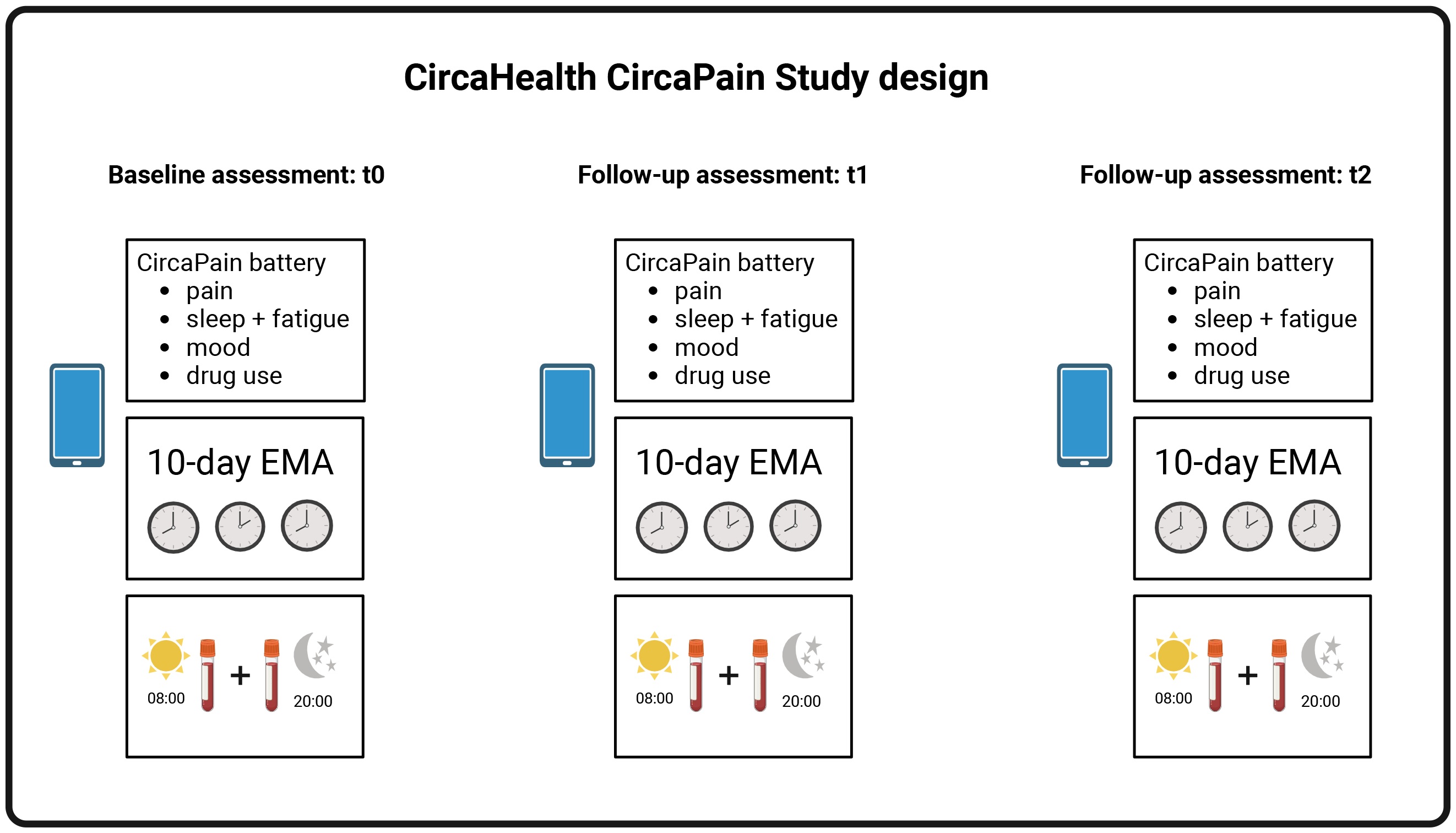
Design and protocol for the CircaHealth CircaPain study. The study is divided into three parts: a baseline assessment (t0) and two follow-up measures at 6 (t1) and 12 months (t2) after the initial assessment. Each assessment includes a battery of questionnaires and a 10-day e-diary, as well as the collection of biological samples.

#### 2.3.1 Patient and Public Involvement

Patient partners (LS and JDC) were involved in the research design from study outset via the CIHR-SPOR Chronic Pain Network. All research questions and outcome measures were developed in collaboration with patient partners, taking into account their priorities and experiences with chronic pain. This was done through consultations with the research team which occur every month. Patient partners contribute to the knowledge translation and dissemination efforts by helping the research team summarise the research objectives and background in clear user-friendly language and ensure that the information is accessible to a public audience.

### 2.4 Participants

#### 2.4.1 Inclusion and exclusion criteria

Individuals with any self-reported chronic pain condition (pain lasting >3 month(30), irrespective of formal diagnosis) who are eligible to participate in the study and complete the survey if: they are at least 18 years old, have internet access, can read, comprehend, and record information written in English or French (or any other languages the survey may be validated in). Individuals reporting a history of travel in the past 2 weeks or planning to travel in a different time zone over the course of the study are excluded to reduce any confounding effects of travel-related circadian disruption (‘jet lag’).

#### 2.4.2 Sample Size and power calculation

We aim to recruit ≥2500 participants within Canada. A subset of participants (n=1000) will be re-contacted for the collection of blood samples to accompany their survey data, to amass 800 participants compliant with epidemiological data and blood collection requirements, allowing for a 20% dropout rate.

Based on our unpublished results, we detect four major pain phenotypes (constant low, constant high, rhythmic, and mixed patterns; at an approximate ratio of ∼1:1:1:1). Mean and standard deviation for each primary variable were obtained from this preliminary data. The sample size calculation used analysis of variance (ANOVA) assuming a type I error *α*= 0.05, and type II error *β*= 0.2 (i.e. 80% power). To detect a difference in pain score and physical function between circadian phenotypes, we will require at least 48 males and 64 females per pain rhythmicity phenotype; to detect changes in rhythmic genes between phenotypes will require at least 20 males and 28 females. We, therefore, collect blood and biopsychosocial data from n=80 males/112 females, ∼200 participants per pain phenotype via online surveys/EMA. Thus, our sample is sufficiently powered to detect significant changes between circadian phenotypes for each sex.

### 2.5 Recruitment procedures

We administer a battery of demographic and clinical self-report questionnaires and daily e-diaries to examine circadian fluctuations in pain. The e-diaries employ an EMA approach which collects repeated measurements of the same participants as they go about their daily lives(31). We collect biological samples (blood) at least 2 times within a ≥ 12-hour period to examine protein profile differences among people with chronic pain. After analysis of location-related data, we aim to expand data collection internationally, validating and translating the survey outside the Canadian population to address latitudinal features.

Passive recruitment strategies are used to reach potential participants. Permission to advertise the study either via social media (Facebook, X, etc.) or in paper form on-site (e.g., poster, handouts) is obtained from interested organizations and will follow their local requirements (e.g., request form, local Research Ethics Board approval, etc.). These could include 1) relevant departments/clinics within partner clinical and community outreach sites (e.g., family health teams, physiotherapy clinics, etc.); 2) specialized pain clinics; or 3) pain advocacy organizations such as PainBC, Association québécoise de la douleur chronique, the People in Pain Network, and the Pain Society of Alberta. Recruitment is also coordinated through partner clinical sites.

Participants can access the survey independently from any device with internet capability. Eligibility screening and informed consent are embedded in the survey and need to be completed before accessing questionnaires. Research team members at partner clinical sites may offer a mobile device (via registered mail) to the participants with Wi-Fi access who do not have a mobile device. Settings on the device will be locked from the user, except for web browsing and Wi-Fi connection access. Currently, validated self-report measures and EMA e-diary are available in English and French. Therefore, the study will be accessible to a majority of Canadians. Furthermore, people who opt-in for blood sample donation will be contacted by the research team and consent after being informed about collection procedures.

To investigate our secondary outcome regarding geophysical location, seasonality, and pain rhythmicity, we will collect information on 1) seasonality via repetition of the study measures (6-12 months follow-ups) and 2) geolocation within Canada and subsequent international expansion (first 3 digits of the postal code). Additional validated translations of questionnaires needed for international expansion will be uploaded to the local Research Ethics Board (REB) for review and approval.

### 2.6 Data collection procedures

Eligible and consented participants are asked to complete questionnaires and e-diaries using their preferred mobile device or home computer, starting the next morning of enrolment. All epidemiological data collection tools are hosted on the platform Research Electronic Data Capture (REDCap)(32, 33), which also facilitates data storage.

#### 2.6.1 Self-report questionnaire battery

Self-report questionnaire battery includes scales/questions covering different domains, based on recommendations from NIH Task Force on reporting chronic low back pain studies(34) and IMMPACT(35, 36): pain and fatigue, affect and other psychological characteristics and physical activity levels (see Table 1). Sociodemographic data and existing medical conditions other than chronic pain are also collected. Participants are asked to complete this questionnaire battery in an online form via a link to the REDCap survey or supported by research staff (see supplementary for English and French versions of the questionnaires). Once completed, participants receive a confirmation email and instructions for the next steps to complete the 10-day e-diary.

#### 2.6.2 10-day e-diary

Using the ecological momentary assessment (EMA) approach(19); the e-diary assesses 7 domains at 3 established timepoints (08:00, 14:00, 20:00) for 10 days. The domains are: 1) pain; 2) fatigue; 3) anxiety; 4) sadness; 5) physical activity; 6) sleep quality and 7) changes in daily medication intake. For pain, participants are asked, “How much pain do you feel right now” (0 = no pain, 10= worst pain imaginable). Similar numerical rating scales or a brief Likert scale are used to assess the other domains (full measures in appendix). A question on sleep quality is included in the 08:00 diary and a question on changes in usual medication intake is included at each time point (see Supplemental Materials for examples of the e-diary monitoring tool in English and French). Invitations and reminders to complete the e-diary are sent either via email or SMS text according to participants’ preference. Notifications are sent respectively at 08:00, 14:00, and 20:00 with a reminder 30 minutes later.

#### 2.6.3 Follow-up measures

Participants will be asked to complete the same measures (self-report questionnaire battery and 10-day e-diary) at 6 and 12 months after initial enrolment to assess seasonality changes. The surveys will be automatically sent to the same account used to complete baseline measures.

#### 2.6.4 Biomarker candidates

##### 2.6.4.1 Biosample collection

Participants can consent to be contacted for the collection of biological samples. Infrastructure for blood collection is under development. Blood samples will be taken from participants across a ≥ 12-hour period, with at least 2 blood draws (e.g., 1 taken in the range 07:00-10:00, 1 taken in the range 19:00-22:00). Once biological sample collection commences, optimal tools (accounting for both feasibility and reliability) will be used and may include the collection of whole blood in K2EDTA and Tempus RNA tubes at collaborating clinical sites or mailable dried blood spot collection kits (e.g., Telimmune DUO Plasma Separation cards, West Lafayette, IN, USA) for participants to complete at home. Biological sample collection methods will be uniform across partner sites and are dependent on available and validated technology at the time of launch. Specific methods for biological sample collection and analysis will be included in future publications.

##### 2.6.4.2 Biosample processing and storage

Depending on available technology at the time of collection, samples will be processed according to best practices associated with individual tools. Blood will be prepared for long-term storage at ≤ -80°C, with the potential for future proteomic, transcriptomic, and/or lipidomic analyses.

##### 2.6.4.3 Bioinformatics

After processing biosamples with the aforementioned analyses, established bioinformatics pipelines will be used. For example, we will evaluate, align, and assemble raw RNA sequencing reads with FastQC(37), Hisat2(38), and StringTie(39), as appropriate. Comparative bioinformatics analysis can then be used to identify genes, transcripts, proteins, and pathways of interest by mining weighted gene coexpression network analysis (WGCNA), as previously done(40-46).

### 2.7 Data management

Any symptom diary containing ≥5 days with submitted diaries at all time points over the 10-day period will be used in the analyses(47); all other symptom diaries will be excluded from main analyses and may be used for secondary analyses. Diary entries completed >1 hour past the time of collection will be excluded from analysis and labelled as ‘timed out’. Any duplicate report for a given report time will be discarded, with only the original dataset used.

### 2.8 Statistics and data analyses

Epidemiological data will be used to identify biopsychosocial determinants of pain and other variables of rhythmicity. Descriptive statistics will be used to characterize the study population. Additionally, the severity of relevant symptomatology reported across sessions will be compared using descriptive and inferential statistics. Our primary outcome is to assess whether there is a change in rhythmicity phenotypes – measured using 10-day e-diary – in both groups from baseline to follow-ups. Clustering of participants based on EMA will be repeated at baseline and follow-up. Participants with distinct rhythmicity trajectories (i.e., pain phenotypes) are identified by EMA scores using a latent class mixed effect model (LCMM, based on *lcmm* R package (48)), a probabilistic modelling algorithm approach that clusters longitudinal data accounting for correlation between repeated measures(49) and has been used to characterize pain chronicity over months(50). To ensure we discover biologically meaningful phenotypes, in addition to LCMM, functional data analysis with high-dimensional data clustering (based on *funHDDC R* package(51)) will be considered as an alternative approach, which allows for participants with similar pain fluctuation phenotypes to be clustered together. Graphical tools (e.g., Sankey plot) will be used to visualize changes in phenotype. Latent transition analysis will be applied to study the probability of transition of an individual from one phenotype to another phenotype from baseline to follow-ups(52, 53). Furthermore, we will use linear mixed-effect models(54, 55) to test for differences in average pain scores in the morning vs evening across time points. All analyses will be performed halfway through and at the end of the study recruitment.

Our secondary outcome is to assess whether changes in rhythmicity phenotypes from baseline to follow-up post-intervention and from different geographical locations (e.g., Eastern vs Pacific time zones, across latitudes) will correspond to changes in biopsychosocial factors measured using our self-report battery. We will use descriptive statistics to characterize the study population (e.g. Fisher’s Exact tests, chi-square, Kruskal-Wallis) where appropriate and we will run regression models to assess any difference between baseline and follow-ups. Once available, blood samples (RNA, protein, etc.) will be analyzed to identify molecular and cellular characteristics of pain rhythmicity.

All statistical analyses will be performed using R(56) or SPSS(57) where appropriate.

## 3. Ethics and dissemination

This study complies with STROBE guidelines(28) and with the principles of the Declaration of Helsinki(29). The CircaHealth CircaPain project and its associated procedures have been reviewed and approved by the Queen’s University Health Sciences and Affiliated Teaching Hospitals Research Ethics Board (DBMS-160-23). Knowledge translation efforts include sharing findings at national and international scientific meetings and online webinars. In addition, we maintain a website (<u>CircaPain</u>) and an X (Twitter) page to post updated resources (blogs, videos, research updates, etc.) and engage with people who live with chronic pain, clinicians, and others interested in the topic. We employ knowledge mobilization in the form of direct data sharing with participants: at the time of consent, participants can opt-in to receive their data upon study completion. They will receive a user-friendly pain report generated using their own data from the 10-day e-diaries after completion of follow-ups to prevent any bias.

## 4. Discussion

This study design adopts the biopsychosocial model of chronic pain through its use of both psychosocial measures and the collection of biological samples for a personalized and precision medicine approach to identifying potential biomarkers of disease. A multidisciplinary approach is adopted to study inter*-* individual variability in pain rhythmicity and its associated psychosocial and molecular factors. We consider fluctuations in pain intensity across time(58), which constitutes an important aspect to take into account in the treatment of chronic pain(21). Indeed, pain intensity is reported to fluctuate over the course of a week and the extent of this variability scores seem to be heterogeneous across participants but stable over assessment periods(59). Recent reviews have shown the importance of several factors when considering pain variability, including weather, seasonality, and genetic variants among others(60, 61). However, there is still an important gap in our knowledge in understanding the causes of this variability(59). New data from preclinical models and analysis of biomarkers in the clinic show a neuro-immune axis in chronic pain. For example, immune cells exhibit rhythmic changes in the blood with increasing numbers during the day peaking in late evening/night(62). Thus, collection of blood samples at different times of day will provide important information regarding rhythmic changes in circulating mediators over time that may affect pain outcomes. Underlying chronobiological and immune mechanisms contributing to pain chronicity will be addressed, providing a crucial aspect in the development of new interventions and preventive measures for chronic pain(10).

The data collected from this longitudinal cohort will identify psychosocial factors and changes in genes/proteins associated with pain rhythmicity in people living with any type of chronic pain. Participants are assessed using a baseline battery of questionnaires and e-diary measures over a 10-day period, which are repeated in follow-up assessments. The baseline assessment employs a modified version of the NIH minimum dataset for research in chronic low back pain (63, 64) to capture key biopsychosocial variables (e.g., pain, fatigue, affect). Furthermore, blood sample collection from this cohort will enable the creation of a chronobiological biobank to analyze the neuroimmune signatures of participants with chronic pain conditions. Any blood samples collected will be banked for future transcriptomic, proteomic, metabolomic and/or lipidomic analysis.

Investigating the impact of pain rhythmicity on psychosocial and neuroimmune outcomes in a clinical population will aid in the identification of novel biopsychosocial markers of chronic pain. Our focus on circadian rhythms as a potential biomarker for pain chronicity represents a novel approach. We consider 1) biopsychosocial factors associated with pain variability and chronobiological rhythmicity; 2) immune cell rhythms and gene expression to identify specific profile and their association with psychosocial outcomes; and 3) seasonality and geophysical location factors that might contribute to pain variability. This interdisciplinary strategy will increase our understanding of mechanisms driving chronic pain and associated psychosocial factors and help identify therapeutic targets acting on the circadian clock or non-pharmacological options that address the circadian control of pain.

## Data Availability

N/a protocol paper

## Author contributions

All authors conceptualized the study. DT was in charge of the study design and wrote the original draft version. DT, HGMG, MGP, NG, and MC were in charge of the inclusion/exclusion criteria and defined how the study would be carried out. DT, HGMG, AMZ, and ZL were in charge of the definition of the data analysis methodology and the statistics, including sample size calculations. LS and JDC contributed to the ethics and dissemination sections. All authors contributed to background research. DT, HGMG, and NG were in charge of the ethics approval process. All authors contributed to the revision of the study. All authors read and approved the final manuscript.

## Acknowledgments

We would like to thank the funding sources for supporting this project.

