## Supplementary material for "The CircaHealth CircaPain study protocol: A longitudinal multi-site study of the chronobiological control of chronic pain": Table 1

**Table 1. Validated and non-standardized measures completed by participants in the CircaPain battery and EMA symptom diaries.** Some questionnaires were not used in their entirety, with relevant subscales/items being isolated.

| **Questionnaire** | **Focus** | **Description** |
| --- | --- | --- |
| *BPI^1^* | Pain and Fatigue | Brief Pain Inventory. Characterizes the nature of a participant’s chronic pain and the degree to which it interferes in their daily activities. Composed of two subscales addressing pain interference in affective factors (e.g., relationships, enjoyment of life, mood) and activity factors (e.g., walking, daily work, general physical activity). |
| *NRS (0-10)* |  | Numerical Rating Scale. An 11-point scale that depicts the severity of pain a participant is feeling at that moment. |
| *PCS-6^2^* |  | Pain Catastrophizing Scale. Assesses participants’ tendency to catastrophize in the face of pain. |
| *PROMIS-29 v2.0^3^* |  | Patient-Reported Outcomes Measurement Information System. Assesses the intensity of pain and fatigue and how these factors impact quality of life. The four-item subscale assessing fatigue is used in the battery. |
| *ISI^4^* | Sleep | Insomnia Severity Index. Assesses severity of insomnia components experienced by a participant. |
| *rMEQ^5^* |  | Reduced Morningness-Eveningness Questionnaire. Assesses alertness of participants and when they feel they are at their “peak” functioning level. |
| *IUS-sf^6 7^* | Affect and other psychological characteristics | Intolerance of Uncertainty Scale (short form). This 12-item scale assesses the emotional and behavioural responses of a participant to the uncertainties experienced in everyday life. |
| *PHQ-4^8^* |  | Patient Health Questionnaire. Briefly assesses for presence of anxious and depressive symptoms among participants along with psychological distress. |
| *POMS^9^* |  | Profile of Mood States. Assesses the mood states of participants. Negative affect items were isolated and used in EMA diary. |
| *GLTEQ^10^* | Physical activity | Godin Leisure-Time Exercise Questionnaire. Assesses a participant’s typical time expenditure on physical activity at different intensity levels. |
| *Non-standardized medical history questionnaire* | Medical history | Records participants’ past and present medical treatments and comorbidities. |

10. The Godin-Shephard leisure-time physical activity questionnaire. *Heal : living well after cancer*;4(1):18.
